# Survival Benefit of Anticoagulation in Patients with End-Stage Kidney Disease and Atrial Fibrillation: A National Population-Based Study

**DOI:** 10.1101/2023.02.09.23285732

**Authors:** Deok-Gie Kim, Sung Hwa Kim, Seong Yong Park, Byoung Geun Han, Jae Seok Kim, Jae Won Yang, Young Jun Park, Jun Young Lee

## Abstract

**Background:** The prevalence of atrial fibrillation (AF) in patients with end-stage kidney disease (ESKD) is high and increasing; however, current evidence is insufficient and conflicting regarding oral anticoagulant (OAC) use in patients with ESKD and AF.

**Methods:** A retrospective cohort study of patients diagnosed with AF after ESKD from January 2007 to December 2017 was conducted using the Korea National Health Insurance System Database. The primary outcome was all-cause death. Secondary outcomes were ischemic stroke, severe bleeding, and major adverse cardiovascular event (MACE). Outcomes were compared between OAC user and nonuser groups using landmark analysis (6 months after AF diagnosis) and 1:3 propensity score matching.

**Results:** Among patients diagnosed with AF after ESKD, the number of patients prescribed any OAC increased 2.3-fold from 2012 (n=3,579) to 2018 (n=8,341), and the proportion prescribed direct OACs (among those prescribed any OAC) increased steadily from 0% in 2012 to 51.4% in 2018. Matched cohort analysis showed that compared with OAC nonusers, OAC users had a lower risk of all-cause death (hazard ratio [HR] 0.71; 95% confidence interval [CI] 0.58–86), ischemic stroke (HR 0.63; 95% CI 0.43–92), and MACE (HR 0.76; 95% CI 0.59–0.98) but no increased risk of severe bleeding (HR 1.58; 95% CI 0.99–2.52). Subgroup analysis according to type of OAC revealed that patients receiving direct OACs had a significantly lower risk of all-cause death (HR 0.44; 95% CI 0.28–0.69), ischemic stroke (HR 0.36; 95% CI 0.13–0.99), and MACE (HR 0.42; 95% CI 0.22–0.79) than OAC nonusers but no increased risk of severe bleeding (HR 0.26; 95% CI 0.04–1.90). Conversely, warfarin use was not associated with all-cause death (HR 0.98; 95% CI 0.79–1.21), ischemic stroke (HR 0.78; 95% CI 0.51–1.19), or MACE (HR 1.03; 95% CI 0.79–1.36) but was associated with a higher risk of severe bleeding (HR 2.43; 95% CI 1.50–3.93).

**Conclusions:** In patients with ESKD and AF, OACs were associated with reduced all-cause death, ischemic stroke, and MACE risks but no increased risk of severe bleeding. Direct OACs were preferable to warfarin with regard to efficacy and safety.

**Clinical Perspective:** *What is new?:*

- Outcomes of oral anticoagulation in patients with end-stage kidney disease and atrial fibrillation are currently unknown.
- In this nationwide cohort study of patients with end-stage kidney disease and atrial fibrillation who had a CHA_2_DS_2_-VASc score of ≥ 1 (men) or ≥ 2 (women), oral anticoagulant therapy was associated with lower risks of all-cause death, stroke, and major adverse cardiovascular event but was no difference in severe bleeding, compared with no anticoagulation.
- When analyzing types of anticoagulants, these reduced risks were found in patients receiving direct oral anticoagulants but not warfarin.

*What are the clinical implications?:*

- Anticoagulation therapy may be superior to no anticoagulation in patients with end-stage kidney disease and atrial fibrillation.
- These findings require confirmation in randomized trials.

## INTRODUCTION

Prevalence rates of nonvalvular atrial fibrillation (AF) in patients with end-stage kidney disease (ESKD) range from 7% to 26% with implantable recorder surveillance, which is much higher than the prevalence of nonvalvular AF in the general population.^1,2^ Additionally, the prevalence of AF in ESKD has been increasing, with a three-fold increase from 3.5% in 1992 to 10.7% in 2006 in patients receiving hemodialysis.^2,3^ Furthermore, among patients with AF, the rates of thromboembolic and bleeding events are higher in patients with ESKD than in those without ESKD.^1^ According to the 2019 American Heart Association/American College of Cardiology/Heart Rhythm Society (AHA/ACC/HRS) guidelines and the 2020 European Society of Cardiology (ESC) guidelines, oral anticoagulants (OACs) are recommended for patients with AF to reduce the risk of stroke and thromboembolic events.^4,5^ For patients with AF who are receiving dialysis, prescribing an OAC is a Class IIb recommendation if the CHA_2_DS_2_-VASc score is 2 or higher in men or 3 or higher in women and a Class IIIb recommendation if the CHA_2_DS_2_-VASc score is 1 in men or 2 in women.^4,5^ Nevertheless, patients with ESKD have an elevated risk of bleeding, mostly related to uremic platelet dysfunction.^6^ Therefore, when deciding whether to prescribe OACs to these patients, it is necessary to consider bleeding complications more carefully than in patients without ESKD. In patients with AF, a meta-analysis of data from several randomized controlled trials (RCTs) showed that warfarin significantly reduced the risk of stroke, compared to placebo or aspirin, but most studies excluded patients with ESKD.^7^ Several retrospective observational studies based on large numbers of patients with ESKD showed that warfarin use increased the risk of stroke and overall mortality in these patients.^8,9^ Therefore, the Kidney Disease: Improving Global Outcome (KDIGO) 2011 consensus statement on cardiovascular disease recommended against routine anticoagulation (warfarin) in patients with ESKD who had AF.^10^ Based on these observational studies and the consensus statement, the rate of warfarin anticoagulation in patients with AF and ESKD with a CHA_2_DS_2_-VASc score ≥ 2 gradually decreased from 2007 to 2013.^11^ A recent meta-analysis of data from 15 unique studies showed that among 47,480 patients with AF and ESKD, only 22.0% were receiving prophylactic warfarin.^12^

Several recent RCTs compared direct oral anticoagulants (DOACs) and warfarin, but most of these studies excluded patients with ESKD.^13-17^ Nevertheless, based on pharmacokinetic data, the United States (U.S.) Food and Drug Administration approved apixaban for use in patients receiving hemodialysis. After this approval, the proportion of DOAC prescriptions among all prescriptions for OACs in patients with ESKD and AF has gradually increased, and as of 2018, the number of patients prescribed DOACs surpassed the number of patients treated with warfarin.^18,19^ A few retrospective cohort studies based on U.S. Renal Data System data and a Korean single-center study showed that OAC use in patients with AF and ESKD was associated with a lower mortality rate, compared to no anticoagulation, although selection bias was a potential limitation of these studies.^20,21^ Several RCTs of OACs in this population are ongoing (e.g., NCT02933697, NCT03987711), but it is very challenging to enroll a sufficient number of participants in these types of studies.^17^ There have been no published RCTs comparing OACs and no anticoagulation in patients with AF and ESKD, and it has not been established whether OACs for AF decrease mortality in patients with ESKD. Therefore, this study aimed to investigate the hypothesis that OACs may reduce the risk of mortality and ischemic stroke without increasing the risk of severe bleeding in patients with ESKD and AF who have a CHA_2_DS_2_-VASc score of ≥ 1 (men) or ≥ 2 (women). To accomplish this, we used nationwide claims-based cohort data that included all patients who underwent renal replacement therapy in South Korea during the study time period.

## METHODS

### Data Source and Study Population

In this study, we analyzed data from the National Health Insurance Service (NHIS) of Korea database. NHIS is a mandatory national health insurance system provided by the Korean government (the Ministry of Health and Welfare). NHIS covers almost the entire (97%) population of the Republic of Korea. Data resources are widely validated and have been used for other studies. NHIS provides data with the approval of NHIS (NHIS-2020-1-467) through the Korean National Health Insurance Sharing Service (http://nhiss.nhis.or.kr). In this study, the requested and approved NHIS data were merged with data from the mortality records database of Statistics Korea (http://mdis.kostat.go.kr), which include the cause and date of mortality. ICD-10 codes were used in the analyses. Details of the specific codes used to define each diagnosis, procedure, and drug in this study are shown in Tables S1 and S2 in the Supplemental Material.

Figure 1 depicts a flowchart describing the study population. We first identified 21,468 patients in the NHIS database who were diagnosed with AF after initiating renal replacement therapy between January 1, 2007 and December 31, 2017. We then excluded 12,485 patients with a contraindication to OAC therapy, such as mitral valve stenosis; those who required an OAC for a non-AF cause, such as systemic embolism, deep vein thrombosis, diagnosed cancer, or post-arthroplasty surgery; and patients with a low stroke risk (CHA_2_DS_2_-VASc score of 0 in men or 0 or 1 in women). As shown in Figure S1 of the Supplemental Material, OAC use was initiated > 6 months after AF diagnosis in almost 40% of patients with ESKD who were prescribed an OAC after being diagnosed with AF. This prolonged time between AF diagnosis and initiation of anticoagulation increases the likelihood that the OAC was prescribed for an indication other than AF; therefore, we used a 6-month landmark analysis to overcome patient selection bias. We also excluded 2,671 patients who developed outcomes (died or were diagnosed with an ischemic stroke) between the cohort entry and landmark dates, were not receiving an OAC consistently (medication possession ratio [MPR] < 80%), or were consistently prescribed an OAC, but the prescription began ≥ 6 months after AF was diagnosed.

**Figure 1.**
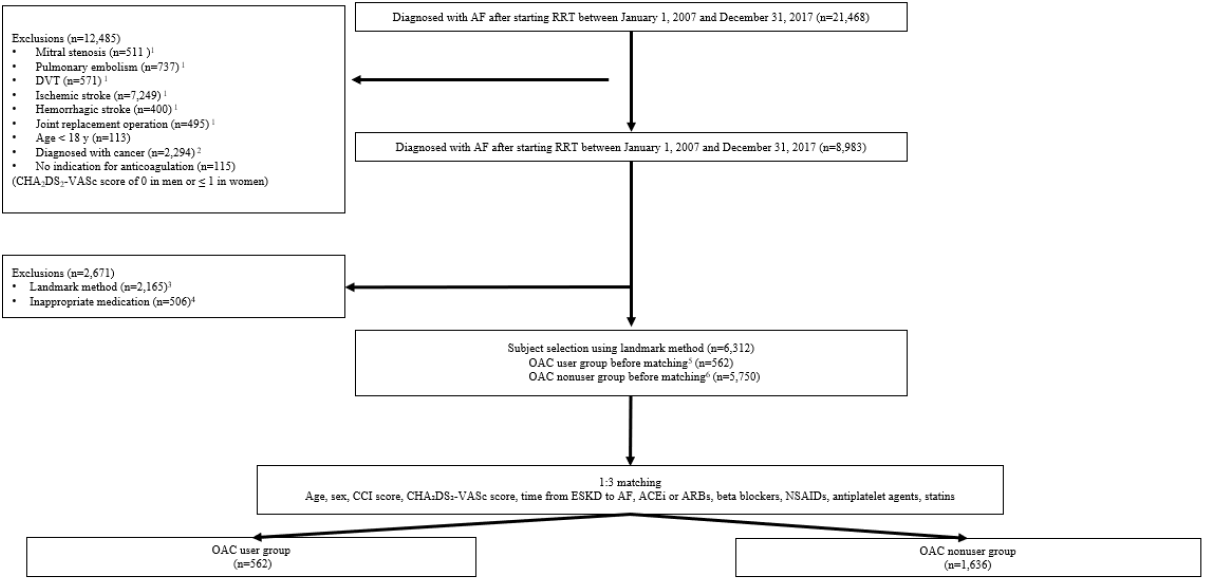
Flow diagram showing selection of study population from the Korean National Health Insurance Service Database. ^1^Diagnosed within 1 year before AF diagnosis. ^2^Diagnosed within 5 years before AF diagnosis. ^3^Patients who died or were diagnosed with an ischemic stroke between the cohort entry and landmark dates were excluded (n=2,165; Figure S2). ^4^Medication possession ratio < 80% (n=119) or prescription for OAC begun > 6 months after AF diagnosis (n=389). ^5^Prescribed an OAC (> 2 prescriptions or total number of prescription days > 30) after AF diagnosis. ^6^Not prescribed any OAC or prescribed only short-duration OAC therapy (< 2 prescriptions or total number of prescription days < 30). Abbreviations: ACEi, angiotensin-converting enzyme inhibitors; AF, atrial fibrillation; ARB, angiotensin receptor blocker; CCI, Charlson Comorbidity Index; DVT, deep vein thrombosis; ESKD, end-stage kidney disease; NSAID, nonsteroidal anti-inflammatory drug; OAC, oral anticoagulant; RRT, renal replacement therapy.

The final OAC user group consisted of 562 patients who were prescribed an OAC after being diagnosed with AF. Specifically, this group included patients who were prescribed an OAC (> 2 prescriptions for OAC or OAC prescribed for a total of > 30 days) within 6 months of the AF diagnosis and whose MPR was above 80%. The initial OAC nonuser group included 5,750 patients with AF who were not prescribed an OAC (either they received no prescription or were prescribed OAC < 2 times or for a total of < 30 days). Patients in this group were then subjected to propensity score matching analysis (as described below), to establish the final matched OAC nonuser group (n=1,636) (Figure 1).

The Institutional Review Board (IRB) of the Yonsei University Wonju College of Medicine (Wonju, Korea) approved this study (IRB number: CR319352), and informed consent was waived because anonymous and de-identified information was used for the analyses. This trial was registered with the Clinical Research Information Service, Republic of Korea (KCT0006397). This work was supported by a National Research Foundation of Korea grant funded by the Korean government (MSIT) (NRF-2020R1G1A1100855). No funder had any role in the study design; data collection, analysis, or reporting; or the decision to submit this study manuscript for publication.

### Propensity Score Matching

We performed propensity score matching in a 1:3 ratio using greedy (nearest neighbor) matching techniques with a caliper of 0.1 standard deviation (SD) to match the OAC user group and OAC nonuser group. Age, sex, Charlson Comorbidity Index (CCI) score, CHA_2_DS_2_-VASc score, time from ESKD diagnosis to AF diagnosis, and medications (e.g., angiotensin-converting enzyme inhibitors [ACEi], angiotensin receptor blockers [ARBs], beta blockers, nonsteroidal anti-inflammatory drugs, antiplatelet agents, and statins) were used to generate propensity scores. After propensity score matching, covariate balance was evaluated by calculating the standardized mean difference of covariates between groups.^22^ These differences were < 0.1 for all covariates, indicative of adequate balance between the matched OAC user and OAC nonuser groups (Figure S2 in the Supplemental Material).

### Data Collection and Study Outcomes

For each patient, we recorded all underlying conditions based on diagnoses reported within 1 year before AF and used these data to calculate the CCI score.^23^ Non-OAC medications that may affect thromboembolic or cardiovascular events were recorded based on prescription information within 3 months before AF diagnosis.

The primary study outcome was all-cause death. The secondary outcomes were the occurrence of ischemic stroke, severe bleeding, and major adverse cardiovascular event (MACE). Severe bleeding was defined as a diagnosis of gastrointestinal bleeding or hemorrhagic stroke requiring hospital admission. MACE was defined as cardiovascular mortality, nonfatal myocardial infarction, or stroke (ischemic or hemorrhagic). The study population was followed until death, 5 years after AF diagnosis, or December 31, 2018, whichever occurred first.

### Statistical Analysis

Baseline characteristics were compared between the OAC user and OAC nonuser groups both before and after matching using the t-test or chi-square test, as appropriate. Categorical variables were expressed as numbers and percentages, and continuous variables were expressed as mean ± SD. Analysis using the landmark approach was performed to reduce immortal time bias. The landmark approach was used to compare the effects of OACs (Figure S3). Kaplan–Meier survival curves and the log-rank test were used to compare the cumulative incidence of outcomes. Hazard ratios (HRs) for each outcome were determined before and after adjusting for age, sex, CHA_2_DS_2_-VASc score, year of ESKD diagnosis, year of AF diagnosis, and time from ESKD diagnosis to AF diagnosis using Cox proportional-hazard regression analysis. For all outcomes except all-cause death, other causes of mortality were considered as competing risks, and regression analyses were performed using Fine and Gray’s model. All p values were two-sided, and p values < 0.05 were considered statistically significant. The statistical analyses were performed using SAS 9.4 (SAS Institute, Cary, NC) and R version 3.63 for Windows (http://cran.r-progect.org/).

## RESULTS

### Oral Anticoagulant Usage Trends and Patient Baseline Characteristics

Among 290,428 patients in South Korea with ESKD between January 1, 2002 and December 31, 2017, 17.6% (n=51,004) were diagnosed with AF (Figure S4 in the Supplemental Material). The number of patients undergoing dialysis who had AF and were receiving OACs increased gradually from 2002 to 2012. Among patients diagnosed with AF after ESKD, the number of patients prescribed any OAC increased 2.3-fold from 2012 (n=3,579) to 2018 (n=8,341). After the release of DOACs in 2012 in Korea, the use of DOACs increased that the proportion prescribed DOACs (among those prescribed any OAC) increased steadily from 0% in 2012 to 51.4% in 2018 (Figure S5 in the Supplemental Material).

After applying the exclusion criteria for the current study (which focused on patients diagnosed with AF after starting renal replacement therapy between January 1, 2007 and December 31, 2017), we found that an OAC was prescribed in only 8.9% (n=562) of the 6,312 patients with ESKD and AF who had a CHA_2_DS_2_-VASc score of ≥ 1 (men) or ≥ 2 (women). Among the 562 patients in our final OAC user group, 337 (60%) were prescribed warfarin, 53 (9.4%) were prescribed apixaban, 98 (17.4%) were prescribed other types of DOACs, and 74 (13.2%) changed OACs during the follow-up period.

Baseline patient characteristics are shown in Table 1. All variables were similar between the 562 patients in the OAC user group and the 1,686 patients in the matched OAC nonuser group. Mean age was 69.3±12.5 years in the OAC user group and 69.4±12.7 years in the OAC nonuser group (p=0.898). Men were more frequent in both groups (57.5% in OAC users vs. 58.2% in OAC nonusers, p=0.767). Mean CHA_2_DS_2_-VASc scores were similar in both groups (3.9±1.7 in OAC users vs. 3.8±1.7 in OAC nonusers, p=0.650), as were CCI scores (4.8±2.3 in OAC users vs. 4.6±2.3 in OAC nonusers, p=0.283). The time from ESKD diagnosis to AF diagnosis was similar between the two groups (921.9±988.3 days in OAC users vs. 890.6±949.3 days in OAC nonusers, p=0.504). Usage of bleeding-related drugs, such as antiplatelet agents, heparin, and NSAIDs, was also similar between groups, as shown in Table 1.

**Table 1.**
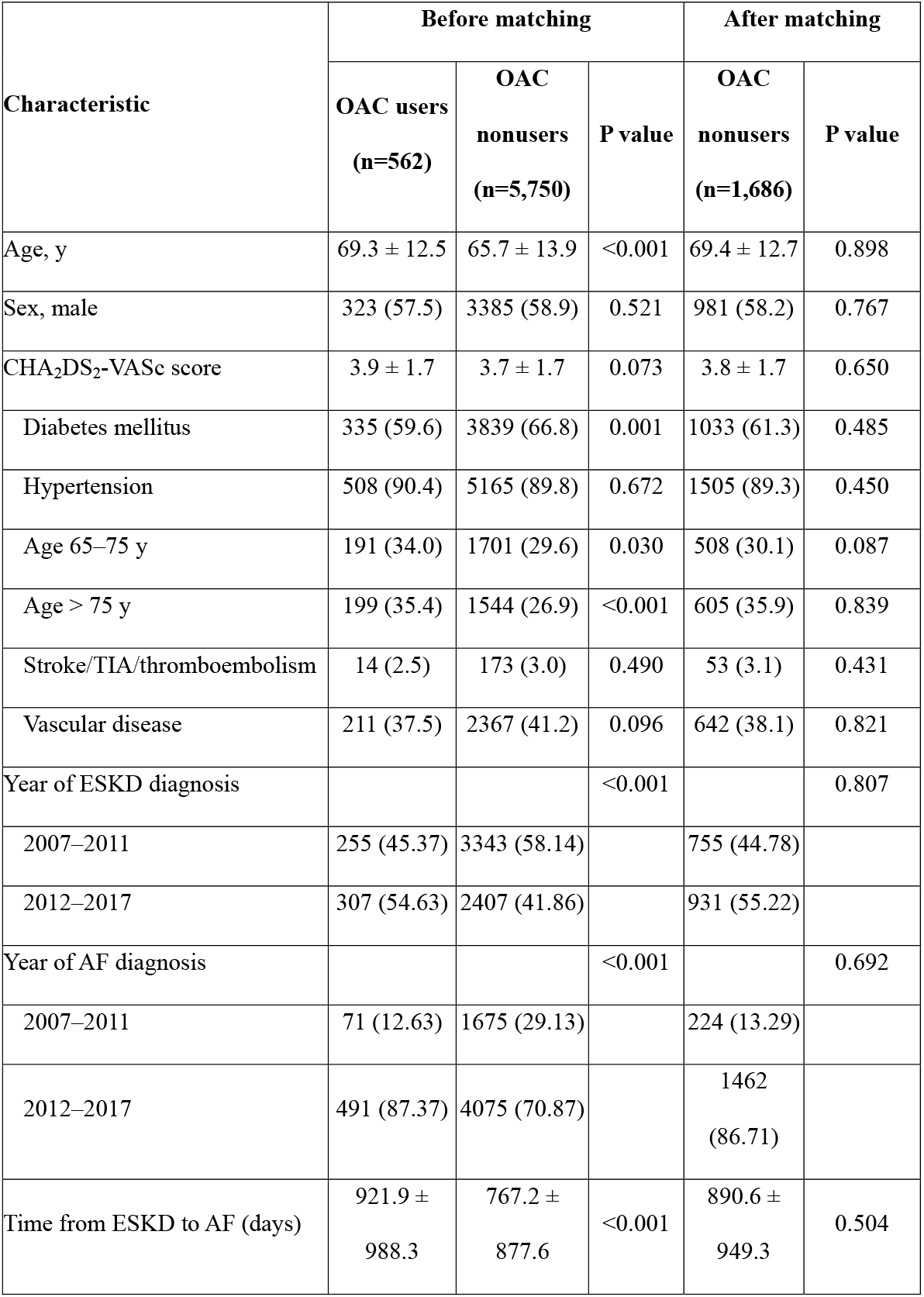
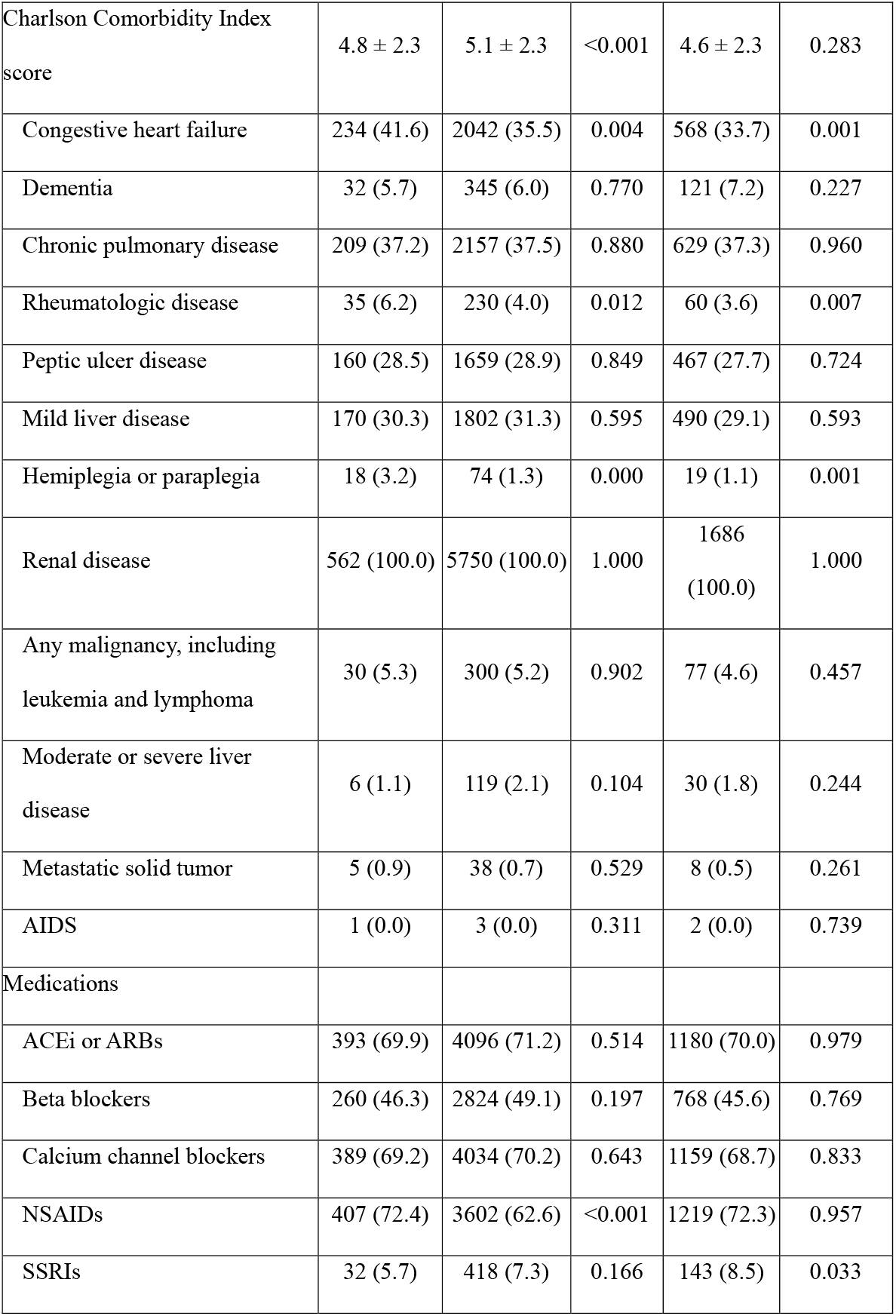
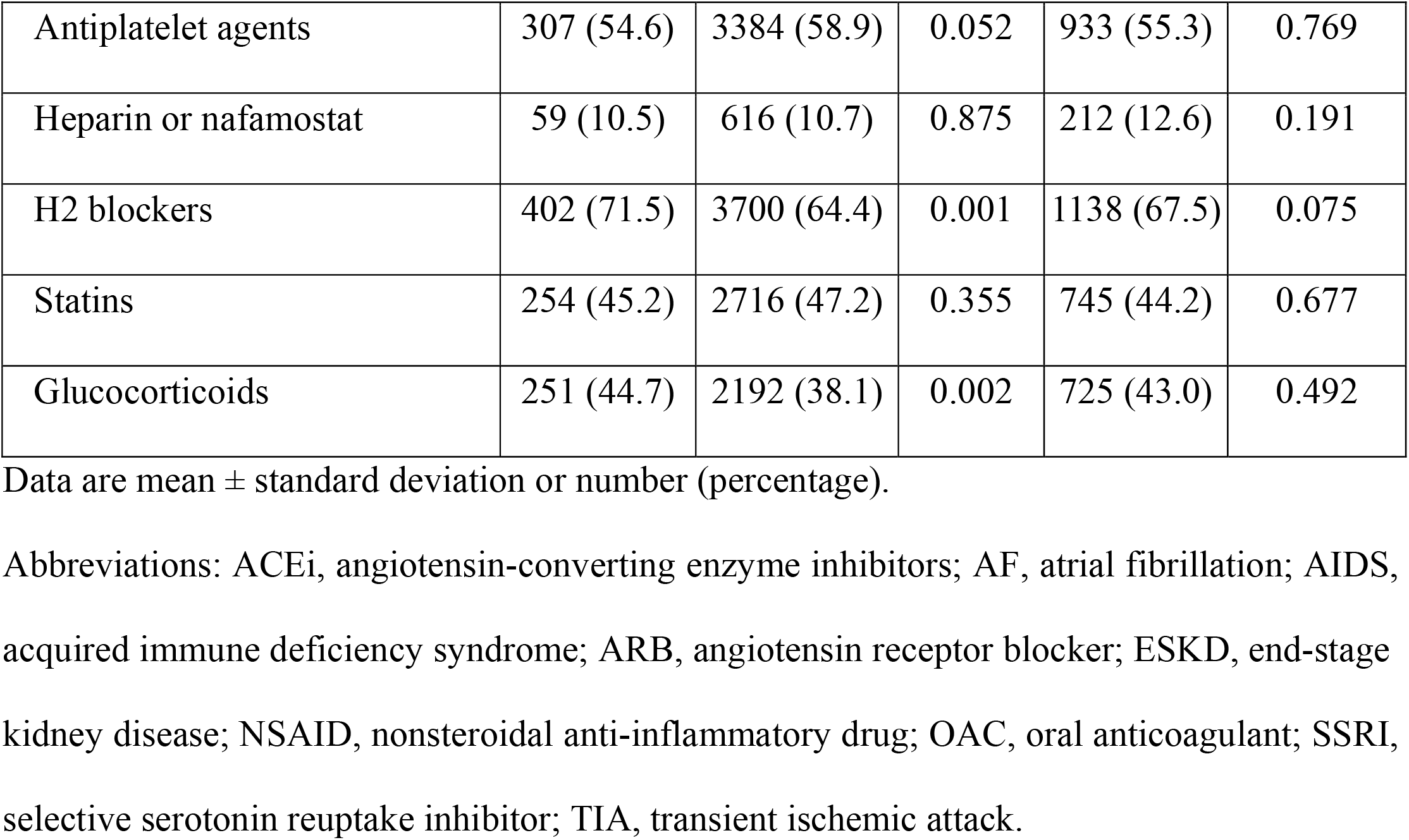
Baseline Patient characteristics.

**Table 2.**
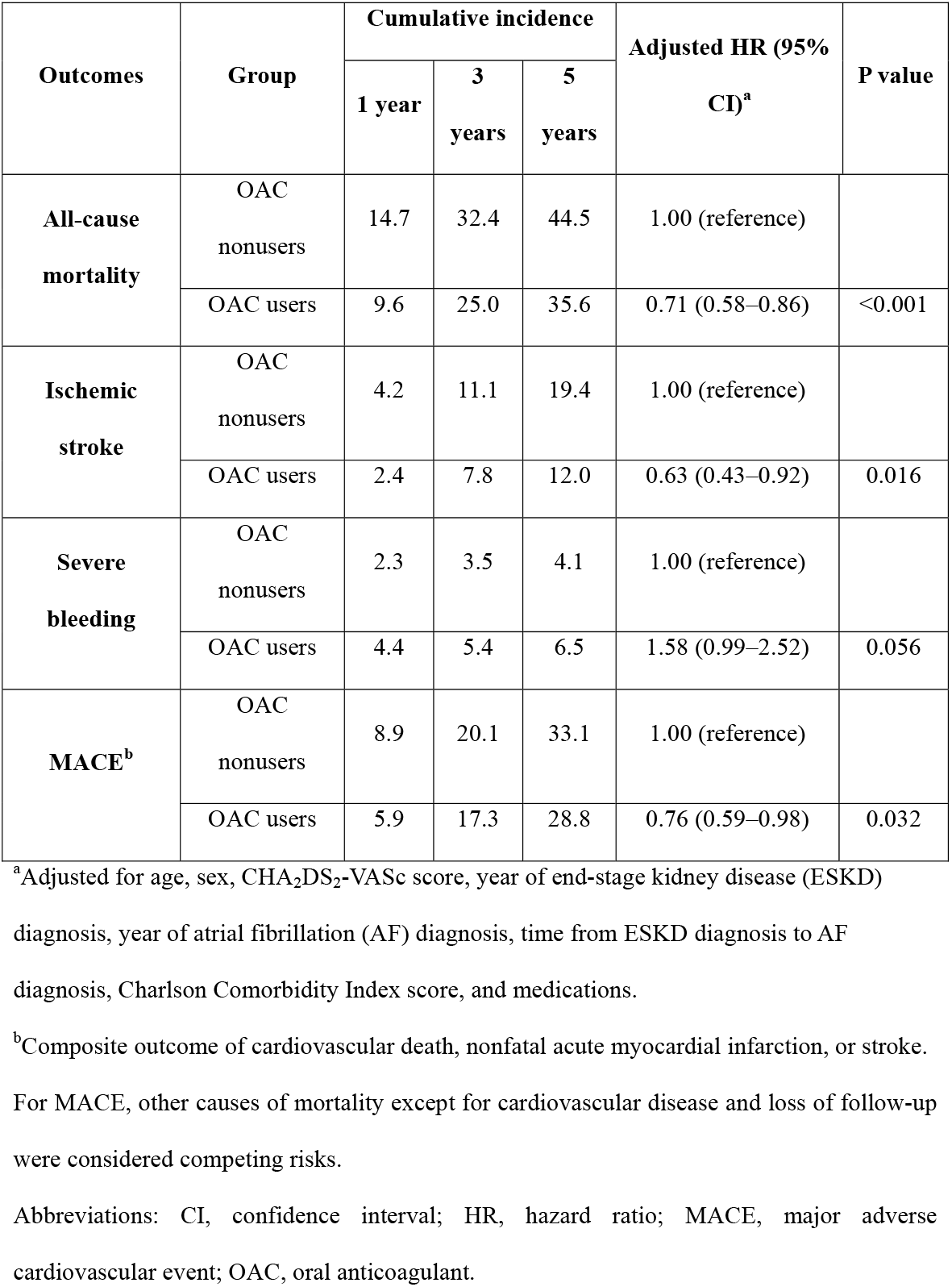
Adjusted hazard ratios for Outcomes According to oral anticoagulant Use.

### Clinical Outcomes

During mean follow-up of 2.65±2.13 years (2.75±2.11 years in OAC users and 2.61±2.13 years in OAC nonusers), 137 (24.4%) patients in the OAC user group and 548 (32.5%) patients in the OAC nonuser group died (p<0.001). In both the OAC user and OAC nonuser groups, cardiovascular disease was the most common cause of death (n=85 [62.0%] in OAC users vs. 291 [53.1%] in OAC nonusers). Other causes of death in both groups are shown in Figure S6 in the Supplemental Material.

In Kaplan–Meier curve analysis, the OAC user group had a significantly lower all-cause mortality than the OAC nonuser group (p<0.001). The cumulative incidences of all-cause death at 1, 3, and 5 years were 9.6%, 25%, and 35.6% in the OAC user group and 14.7%, 32.4%, and 44.5% in the OAC nonuser group. The multivariable adjusted HR for all-cause death in OAC users (compared to nonusers) was 0.71 (95% confidence interval [CI] 0.58– 0.86, p<0.001). The probability of ischemic stroke and MACE were also significantly lower in the OAC user group than in the OAC nonuser group (Figure 2). The multivariable adjusted HRs for ischemic stroke and MACE were 0.63 (95% CI 0.43–0.92, p=0.016) and 0.76 (95% CI 0.59–0.98, p=0.032) in OAC users, compared to OAC nonusers. The probability of severe bleeding was significantly higher in the OAC user group than in the OAC nonuser group (p=0.042). The cumulative incidences of severe bleeding at 1, 3, and 5 years were 4.4%, 5.4%, and 6.5% in the OAC user group and 2.3%, 3.5%, and 4.1% in the OAC nonuser group. However, OAC use was not associated with an increased risk of severe bleeding in the multivariable adjusted analysis (HR 1.58, 95% CI 0.99–2.52, p=0.056).

**Figure 2.**
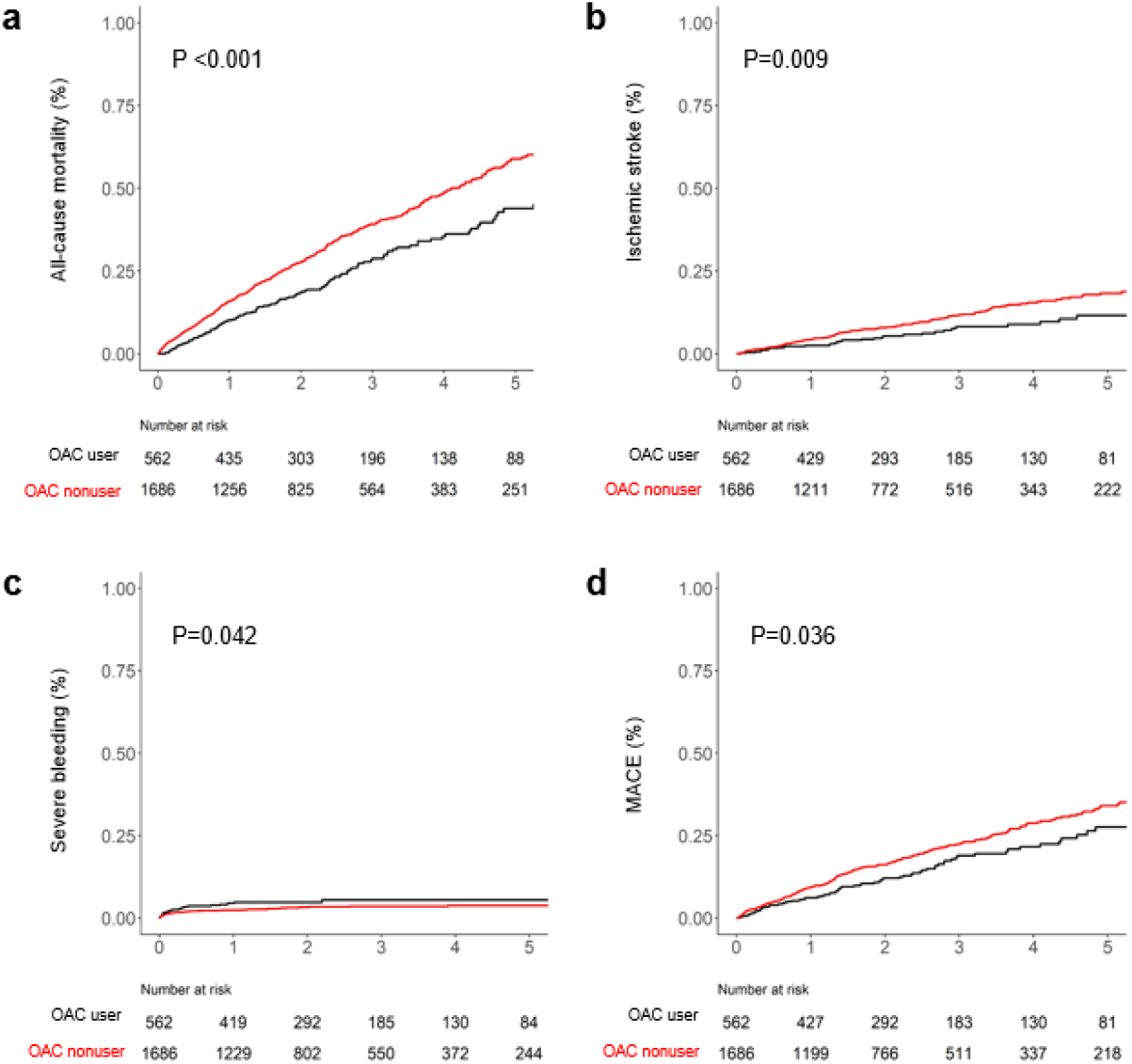
Kaplan–Meier curve analyses for each outcome. a. All-cause mortality, b. ischemic stroke, c. severe bleeding, and d. MACE. MACE is the composite outcome of cardiovascular mortality, nonfatal myocardial infarction, or stroke. Abbreviations: MACE, major adverse cardiovascular event; OAC, oral anticoagulant.

### Subgroup Analysis

The results of subgroup analysis according to patient characteristics are shown in Figure S3 in the Supplemental Material. OAC use was associated with a mortality benefit in patients receiving hemodialysis (HR 0.71, 95% CI 0.58–0.86), elderly patients (age ≥ 65 years) (HR 0.65, 95% CI 0.52–0.81), and patients with a CHA_2_DS_2_-VASc score ≥ 2 (men) or ≥ 3 (women) (HR 0.69, 95% CI 0.57–0.85). OAC use was also associated with protective effects for ischemic stroke and MACE in patients receiving hemodialysis (HR 0.63, 95% CI 0.43– 0.92 and HR 0.77, 95% CI 0.60–0.99, respectively) and patients with a CHA_2_DS_2_-VASc score ≥ 2 (men) or ≥ 3 (women) (HR 0.60, 95% CI 0.41–0.89 and HR 0.72, 95% CI 0.56– 0.94, respectively). OAC was associated with an increased risk of severe bleeding in patients undergoing hemodialysis (HR 1.63, 95% CI 1.02–2.61) and elderly patients (age > 80 years) (HR 3.63, 95% CI 1.31–10.09).

In subgroup analysis according to type of OAC, warfarin was associated with an increased risk of severe bleeding (HR 2.43, 95% CI 1.50–3.93), with no reduction in risk of mortality, ischemic stroke, or MACE. In contrast, DOACs were associated with a reduced risk of death (HR 0.44, 95% CI 0.28–0.69), ischemic stroke (HR 0.36, 95% CI 0.13–0.99), and MACE (HR 0.42, 95% CI 0.22–0.79) and no increased risk of severe bleeding (HR 0.26, 95% CI 0.04–1.90) (Figure 3).

**Figure 3.**
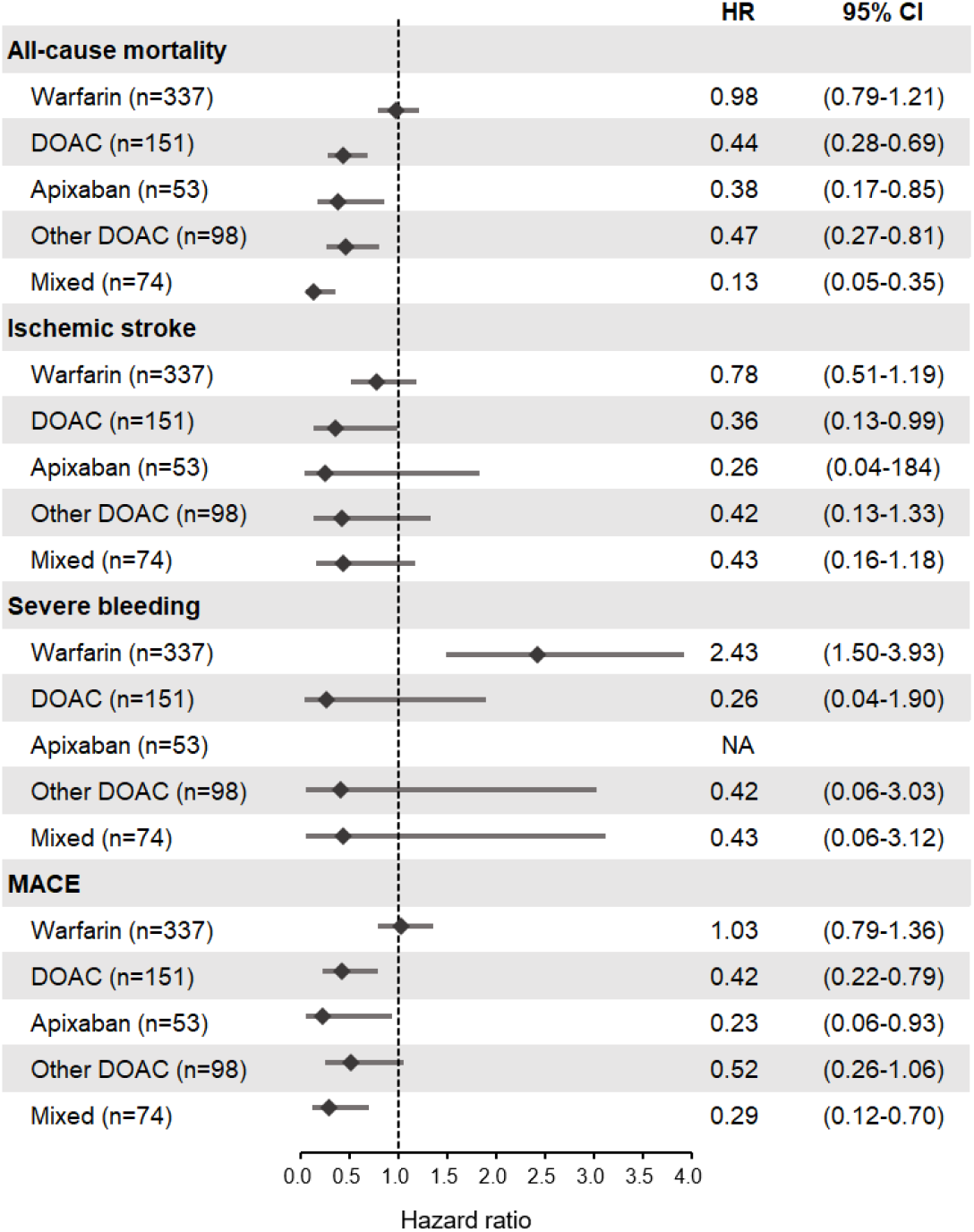
Subgroup analysis for all-cause mortality, ischemic stroke, severe bleeding, and MACE according to type of oral anticoagulant. Abbreviations: CI, confidence interval; DOAC, direct oral anticoagulant; HR, hazard ratio; MACE, major adverse cardiovascular event.

## DISCUSSION

This nationwide cohort study showed that among patients with ESKD and AF (n=51,004), 42.1% (n=21,468) were diagnosed with AF after starting renal replacement therapy. Of the 21,468 patients with ESKD and AF, only 2,198 (OAC users and nonusers) were eligible for inclusion in this “real world” cohort study after applying the study exclusion criteria. We found that compared to no anticoagulant therapy, appropriate anticoagulant therapy was associated with a reduction in all-cause death, ischemic stroke, and MACE, without increasing the risk of severe bleeding. Among anticoagulants, warfarin was associated with an increased risk of severe bleeding without benefits, whereas DOACs were associated with a decreased risk of all-cause death, ischemic stroke, and MACE, without increasing the risk of severe bleeding.

There are conflicting data regarding the benefits and risks of OAC use in patients with ESKD who have AF. U.S. Medicare claims data analysis showed no association between OACs and the risk of mortality or stroke.^11^ Although both warfarin and DOACs are now available for patients with ESKD and AF,^4,5^ warfarin was the only OAC option for decades. Despite extensive experience with warfarin in patients with ESKD, the amount of time patients are within the target international normalized ratio (INR) range for warfarin is low, even in clinical research settings. In a retrospective study, INR was within the target therapeutic range in only 21% of patients with AF and ESKD treated with warfarin.^17,24,25^ In addition to increasing the risk of bleeding, warfarin may also trigger calciphylaxis, a vasculopathy most common in patients with ESKD.^26^ Several studies have shown that warfarin usage in patients with ESKD and AF is associated with a higher risk of hemorrhagic stroke and no change in all-cause death or ischemic stroke.^11,12,27^ One study from the Netherlands showed that compared to no anticoagulation therapy, warfarin increased the risk of all-cause death; however, 26.4% of patients in that study had a CHA_2_DS_2_-VASc score < 2.^28^ In contrast, a nationwide Danish registry study showed that warfarin was associated with a lower risk of death in patients with ESKD and AF who had a CHA_2_DS_2_-VASc score ≥ 2.^29^ However, that study did not evaluate the risk of severe bleeding, only the risk of any stroke or any bleeding. Our results showed that warfarin did increase the risk of severe bleeding. Therefore, when using warfarin for anticoagulant therapy, the risk of bleeding must be taken into account, and several strategies must be considered to try to reduce this risk, such as monitoring the INR frequently and using gastrointestinal protective medications. Patients receiving hemodialysis are especially susceptible to bleeding, independent of OAC use, because heparin therapy is required to maintain the patency of the dialysis pathway (arteriovenous fistula or hemodialysis catheter).

We found that the proportion of warfarin prescriptions among all anticoagulant prescriptions in patients with ESKD and AF gradually decreased after 2012, and by 2018, the number of prescriptions for DOAC surpassed the number of warfarin prescriptions, which is similar to the U.S. national cohort study results.^18^ Several retrospective cohort studies compared the effects of warfarin versus DOAC in patients with ESKD and AF. A Taiwanese nationwide retrospective cohort study showed no significant difference in risk of ischemic stroke, systemic embolism, or major bleeding between DOAC and warfarin in this patient population.^30^ Retrospective cohort studies from the U.S. Renal Data System showed that for patients with ESKD and nonvalvular AF, apixaban was associated with a lower risk of major bleeding but no difference in the risk of systemic embolism or stroke, compared to warfarin.^18,27,31^ In patients with ESKD and nonvalvular AF, dabigatran was associated with an increased risk of major bleeding but no difference in risk of stroke or systemic embolism, compared to warfarin.^19,27^ Also in this patient population, some studies demonstrated that rivaroxaban was associated with a similar or lower risk of major bleeding or thromboembolism, compared to warfarin,^32,33^ whereas a multicenter RCT showed that it reduced both cardiovascular events and major bleeding, compared to warfarin.^34^ A retrospective study of patients with nonvalvular AF and ESKD reported no significant differences between rivaroxaban and apixaban for any effectiveness or safety outcomes.^35^ A U.S. study showed that compared to no anticoagulation, apixaban was associated with a higher incidence of major bleeding, without a lower incidence of stroke or systemic thromboembolism.^20^ However, other studies, including a meta-analysis, showed that apixaban was associated with a lower risk of major bleeding than warfarin.^18,27,31^ Therefore, apixaban and rivaroxaban may be preferred over warfarin in patients with ESKD and AF.

Compared to patients with AF who have normal kidney function, the risk of stroke is more than five-fold higher in patients with AF and ESKD.^29^ Furthermore, the risk of death in patients with ESKD is much higher in those with AF, compared to those without AF.^3^ Therefore, studies of the optimal way to reduce the risk of ischemic stroke and systemic embolism through appropriate anticoagulation therapy are warranted. As our results suggest that anticoagulation is associated with a reduced risk of all-cause death through its beneficial effects on ischemic stroke, initiating appropriate and individualized anticoagulation therapy to prevent ischemic stroke may ultimately improve survival in patients with ESKD and AF.

Because patients with ESKD have many comorbidities, it is difficult to select similar patients when conducting a comparison study. A recent RCT also failed to recruit a sufficient number of patients.^17^ Even in our current nationwide cohort study, only 2,198 of 299,084 patients with ESKD were eligible for inclusion in the study. In contrast to other cohort studies, our study used several strategies to overcome bias, which is an inherent limitation of retrospective observational studies. In addition to landmark analysis and propensity score matching, by analyzing only those patients who were diagnosed with AF after being diagnosed with ESKD, we could set the date of the first AF diagnosis as the index date, thereby reducing lead time bias due to AF duration.^36^ Also, in contrast to most other researchers, we used the most recent updated CHA_2_DS_2_-VASc score for patient inclusion criteria and sex-specific cut-off values for recommended OAC use.^4,5^ We observed beneficial effects in patients with higher CHA_2_DS_2_-VASc scores, consistent with the results of a Danish study, which also found that OACs were more beneficial in patients with ESKD who had a higher CHA_2_DS_2_-VASc score.^29^

Nevertheless, this study has some limitations. Because of the limited information included in the claims database, we could not obtain detailed patient information, such as laboratory results or the dose of OAC. We, therefore, could not completely eliminate selection bias related to these parameters. However, our study showed how OACs are used in real-world practice, and use of warfarin is inherently limited by an inability to reliably achieve target INR values. In addition, the diagnoses of cardiovascular and cerebrovascular disease were established through operational definitions, which may have led to misdiagnosis.

In conclusion, this study showed that in patients with nonvalvular AF and ESKD who had a CHA_2_DS_2_-VASc score ≥ 1 (men) or ≥ 2 (women), OAC therapy was associated with a lower risk of death, MACE, and ischemic stroke. However, because of the nature of ESKD, anticoagulants may be especially likely to increase the risk of bleeding in this patient population. Therefore, individualized anticoagulant therapy should be considered in these patients to reduce the likelihood of major bleeding.

## Data Availability

All data is available through approval of National Health Insurance Service Korea.

https://nhiss.nhis.or.kr/bd/ay/bdaya001iv.do

## Nonstandard Abbreviations and Acronyms

OAC: oral anticoagulant
NHIS: National Health Insurance Service

## Acknowledgments

None.

## Supplemental Materials

Table S1-S3 Figure S1-S6

